# Cohort profile: Virus Watch: Understanding community incidence, symptom profiles, and transmission of COVID-19 in relation to population movement and behaviour

**DOI:** 10.1101/2023.01.31.23285232

**Authors:** Thomas Byrne, Jana Kovar, Sarah Beale, Isobel Braithwaite, Ellen Fragaszy, Wing Lam Erica Fong, Cyril Geismar, Susan Hoskins, Annalan M D Navaratnam, Vincent Nguyen, Parth Patel, Madhumita Shrotri, Alexei Yavlinsky, Pia Hardelid, Linda Wijlaars, Eleni Nastouli, Moira Spyer, Anna Aryee, Ingemar Cox, Vasileios Lampos, Rachel A McKendry, Tao Cheng, Anne M Johnson, Susan Michie, Jo Gibbs, Richard Gilson, Alison Rodger, Ibrahim Abubakar, Andrew Hayward, Robert W Aldridge

**Affiliations:** Centre for Public Health Data Science, Institute of Health Informatics, University College London, UK; Institute of Epidemiology and Health Care, University College London, London, UK; Department of Infectious Disease Epidemiology, London School of Hygiene and Tropical Medicine, Keppel Street, London, UK; MRC Centre for Global Infectious Disease Analysis, Department of Infectious Disease Epidemiology, School of Public Health, Imperial College London, London, UK; UCL Great Ormond Street Institute of Child Health, London, UK; Department of Population, Policy and Practice, UCL Great Ormond Street Institute of Child Health, London, UK; Francis Crick Institute, London, UK; University College London Hospital, London, United Kingdom; Department of Computer Science, University College London, London, UK; London Centre for Nanotechnology and Division of Medicine, London, UCL; SpaceTimeLab, Department of Civil, Environmental and Geomatic Engineering, University College London, London, UK; Centre for Population Research in Sexual Health and HIV, Institute for Global Health, London, UK; Centre for Behaviour Change, University College London, London, UK; Institute for Global Health, University College London, London, UK; Royal Free London NHS Foundation Trust, London, UK

## Abstract

**Key Features:**

- Virus Watch is a national community cohort study of COVID-19 in households in England and Wales, established in June 2020. The study aims to provide evidence on which public health approaches are most effective in reducing transmission, and investigate community incidence, symptoms, and transmission of COVID-19 in relation to population movement and behaviours.
- 28,527 households and 58,628 participants of age (0-98 years, mean age 48), were recruited between June 2020 - July 2022
- Data collected include demographics, details on occupation, co-morbidities, medications, and infection-prevention behaviours. Households are followed up weekly with illness surveys capturing symptoms and their severity, activities in the week prior to symptom onset and any COVID-19 test results. Monthly surveys capture household finance, employment, mental health, access to healthcare, vaccination uptake, activities and contacts. Data have been linked to Hospital Episode Statistics (HES), inpatient and critical care episodes, outpatient visits, emergency care contacts, mortality, virology testing and vaccination data held by NHS Digital.
- Nested within Virus Watch are a serology & PCR cohort study (n=12,877) and a vaccine evaluation study (n=19,555).
- Study data are deposited in the Office of National Statistics (ONS) Secure Research Service (SRS). Survey data are available under restricted access upon request to ONS SRS.

## Why was the cohort set up?

The United Kingdom Research and Innovation (UKRI) Medical Research Council (MRC) & the Department of Health and Social Care National Institute for Health and Care Research (DHSC NIHR) funded Virus Watch in April 2020 under the COVID-19 Rapid Response Call 2. During the early stage of the pandemic in the UK, (February/March 2020), data on COVID-19 was largely collected in hospital settings. Our aim was to bring together an experienced team of respiratory infectious disease epidemiologists to rapidly establish a national community cohort study of COVID-19 in households living in England and Wales that built upon our experience from Flu Watch - a community cohort designed to estimate community burden of influenza and influenza-like illness (1). The DHSC/UKRI awarded additional funding to the study (under the Rapid Response Initiative call) in August 2020 to recruit larger numbers of minority ethnic and migrant populations when it became increasingly apparent that these groups were underrepresented in research studies whilst experiencing greater risk of hospitalisation and death from COVID-19.

Virus Watch aims to provide evidence on which public health approaches are most likely to be effective in reducing the spread and impact of the virus and investigates community incidence, symptom profiles, and transmission of COVID-19 in relation to population movement and behaviour (2).

The main research questions we set out to address were: what is the rate of infection; what is the rate of infected people experiencing symptoms; what is the rate of people seeking healthcare; what are the hospitalisation and mortality rates associated with COVID-19, at different points in time and within different population groups. We wanted to describe COVID-19 symptoms and their severity and to understand people ‘s behaviour in terms of infection prevention as well as movement, travel, activities and social contact. Understanding how these outcomes differ by ethnicity, migration and deprivation, and what risk factors may explain any differences, constituted some of our key objectives.

In addition, we wanted to understand how negative consequences of the COVID-19 pandemic and public health control measures affect economic circumstances and mental health. We had a particular focus on people from minority ethnic and migrant backgrounds and how access to primary care for COVID-19 varied among these groups compared to the White British population and what factors might explain this.

After vaccines became available in the UK in December 2020, further funding was provided by the DHSC for undertaking serological testing for a subset of the Virus Watch cohort. This program ran from February 2021 – April 2022 and was designed to assess the effectiveness and impact of COVID-19 vaccines on both symptomatic and asymptomatic infections and on transmission. We wanted to compare the effectiveness of vaccines in different population subgroups and assess the duration of protection, correlates of protection and immunity against emerging strains.

## Who is in the cohort?

As of the 7th of July 2022, 58,628 participants aged 0 - 98 years (mean age 48 years) from 28,527 households had enrolled into Virus Watch (see **Table 1**).

**Table 1.** Demographics of Virus Watch study participants

| Characteristic | Virus Watch Participants | ONS (%)* |
| --- | --- | --- |
| **All** | **58,628** | **–** |
| **Age group (years)** |  |  |
| 0-15 | 7,371 (13%) | 19% |
| 16-24 | 3,499 (6.0%) | 11% |
| 25-44 | 11,725 (20%) | 26% |
| 45-64 | 19,657 (34%) | 26% |
| 65+ | 16,376 (28%) | 19% |
| **Sex (self-reported)\*\*** |  |  |
| Male | 21,687 (37%) | 49% |
| Female | 27,506 (47%) | 51% |
| Other/Missing/Prefer not to say | 9,435 (16%) | — |
| **Sex (including derived)\*\*** |  |  |
| Male | 26,274 (45%) | 49% |
| Female | 31,533 (54%) | 51% |
| Other/Missing/Prefer not to say | 821 (1.4%) | — |
| **Ethnicity** |  |  |
| White British | 40,481 (69%) | 81% |
| White Irish | 671 (1.1%) | 1% |
| White Other | 2,816 (4.8%) | 4% |
| Mixed | 998 (1.7%) | 2% |
| South Asian | 2,728 (4.7%) | 5% |
| Other Asian | 427 (0.7%) | 2% |
| Black | 493 (0.8%) | 3% |
| Other Ethnicity | 288 (0.5%) | 1% |
| Prefer not to say | 192 (0.3%) | — |
| Missing | 9,534 (16%) | — |
| **Region** |  |  |
| North East | 2,528 (4.3%) | 5% |
| North West | 5,572 (9.5%) | 12% |
| Yorkshire and The Humber | 3,035 (5.2%) | 9% |
| East Midlands | 4,945 (8.4%) | 8% |
| West Midlands | 3,020 (5.2%) | 10% |
| East of England | 10,545 (18%) | 11% |
| London | 9,083 (15%) | 15% |
| South East | 9,845 (17%) | 15% |
| South West | 3,956 (6.7%) | 10% |
| Wales | 1,532 (2.6%) | 5% |
| Missing | 4,567 (7.8%) | — |

\*ONS = Office for National Statistics.
ONS data for age sex and region drawn from Mid-2019
Estimates of the Population for the UK, England, and Wales, Scotland and Northern Ireland
(figures for England and Wales) (3)
ONS data for ethnicity drawn from 2011 Census (figures for England and Wales) (4)
\*\* Sex at birth was self-reported. If missing, sex was obtained via data linkage or derived via name-gender matching
based on US names from 1930-2015 https://data.world/howarder/gender-by-name (5)

Virus Watch is a prospective household community cohort study. Recruitment methodology was adapted throughout the study (see **Table S1**) to ensure the target number of participants from the general population was reached. We also aimed to recruit a sufficiently large sample of participants from minority ethnic backgrounds to investigate infection risk and impact of the pandemic on specific groups of interest. Postcards, leaflets and adverts used to recruit participants were designed to inform individuals about the study and direct them to our website http://ucl-virus-watch.net/ where they could self-select into the study.

We used the Royal Mail Post Office Address File to generate a list of sampled residential addresses to send Virus Watch recruitment postcards to. The initial sample design was a single-stage stratified probability sample. Within each region, residential addresses were sorted by quintiles of Index of Multiple Deprivation (IMD), within quintiles by local authorities, postcodes and address (2). We also delivered invitation leaflets to letterboxes in residential areas around our blood taking clinics to ensure we could recruit our target number of participants into the laboratory subcohort. Enrolled participants were invited to join the laboratory subcohort if they lived within 5km (urban areas) or 10km radius (rural areas) of the nearest blood taking clinic for serological sampling.

We worked with nine of the 15 NIHR Local Clinical Research Networks (LCRNs) across England to send SMS messages from General Practitioner (GP) clinics to their patient lists with a link to the study website inviting participants to take part. To boost recruitment of minority ethnic participants into the cohort, we sent targeted letters based on ethnicity via 90 GP clinics from 9 LCRNs that included a £20 voucher incentive per household to sign up.

Digital invitations were shared via trusted networks including patient advocacy group websites, Twitter, Facebook and WhatsApp. We also undertook a paid advertising campaign via Facebook.

Study participants were emailed and asked to share an invite with their family and friends. Throughout the pandemic, multiple newspaper articles, radio and TV appearances of the study team also contributed to public-facing exposure and recruitment.

## How often have the participants been followed up?

After signing up to the study and completing a baseline survey for every member of the household, a nominated household study lead completed a weekly online illness survey, and monthly surveys (from Dec 2020) about pandemic-relevant sociodemographic and clinical factors.

Participants in the laboratory subcohort were invited to either attend a clinic in their local area or schedule a home visit to have their blood taken on 2 occasions (Oct 2020 - Jan 2021 and again between May - Aug 2021). Participants that were unable to visit a clinic and could not receive a home visit were asked to provide a finger prick sample (in clinic or self-collected at home). Between Oct 2020 and May 2021, participants also posted self-administered nasal swabs for PCR assays of SARS-CoV-2 if they experienced any of the following symptoms for 2 or more days; fever, cough or loss or change of sense of taste or smell. The design of the laboratory subcohort, including the sample collection algorithm and specific symptoms of interest has been published in the study protocol (2).

Adults taking part in the vaccine evaluation subcohort posted self-collected finger prick capillary blood microsamples monthly between Feb-Aug 2021, and every other month from Sep 2021-Mar 2022.

For participants living in England, linkage of Virus Watch to data held by NHS Digital (Hospital Episode Statistics, Death Registrations, National Immunisation Management System (NIMS), COVID Vaccine Adverse Events Data, virological surveillance data) will take place quarterly during the study and for up to 5 years after the end of the study (until 30 September 2026).

Retention of participants in the study has decreased over time. Many participants dropped out of the study and failed to complete any weekly surveys after enrolling but among those who completed at least one weekly survey, engagement was high. In the first 6 months of the study approximately 75% of enrolled participants were regularly completing the weekly illness surveys (**Figure 2**). Over the course of the pandemic the proportion of participants who were lost to follow-up steadily increased and by May 2022 the proportion still regularly completing surveys had reduced to around 50% of enrolled participants. Participants self-select into the study and are free to stop participating at any time. We used a 75% survey completion rate for weekly surveys as a cut-off to compare the characteristics of high responders and low responders. Participants who completed less than 75% of all possible weekly surveys were more likely to be younger, from an ethnic minority background and living in London (Table 2).

**Table 2:**
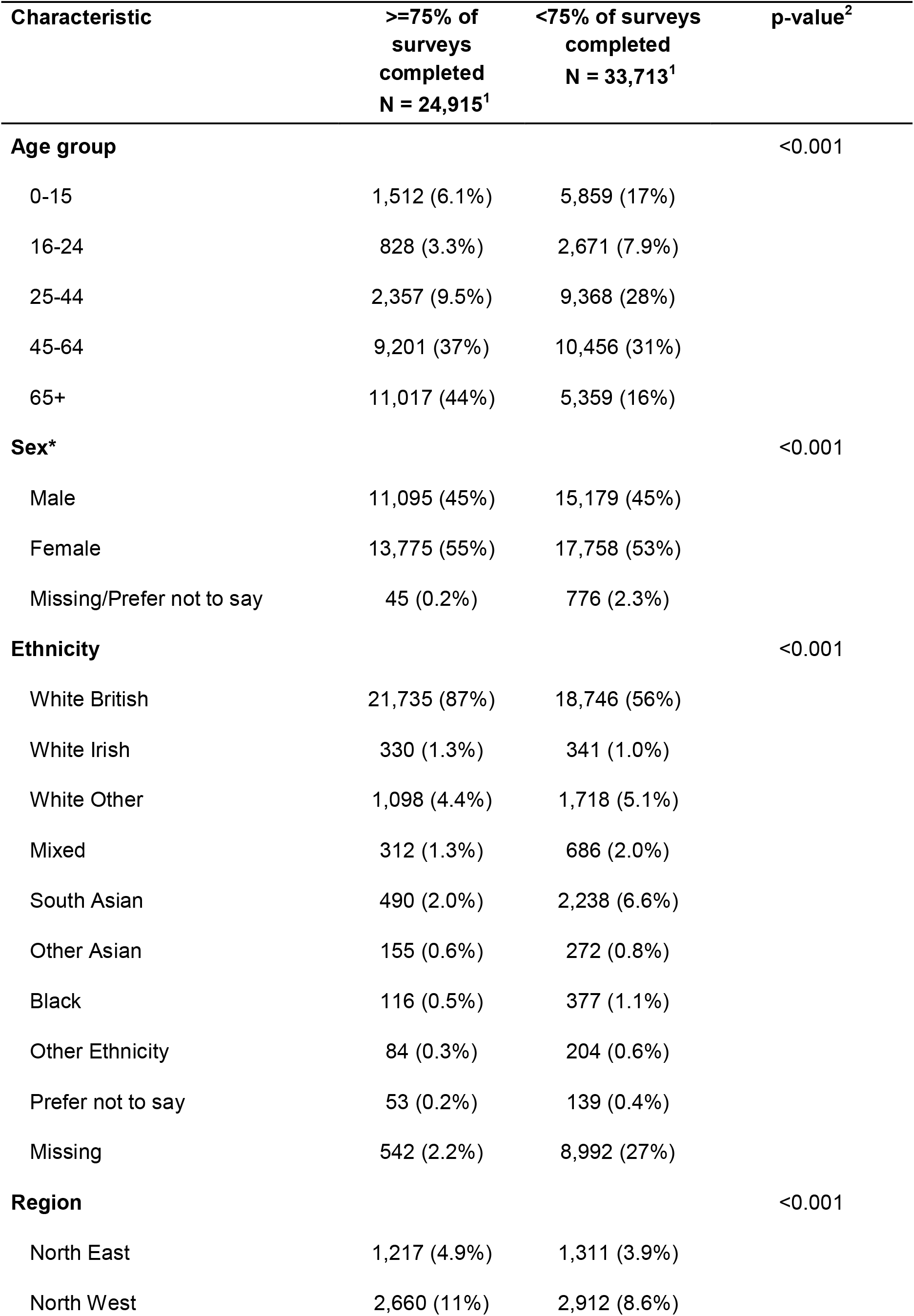
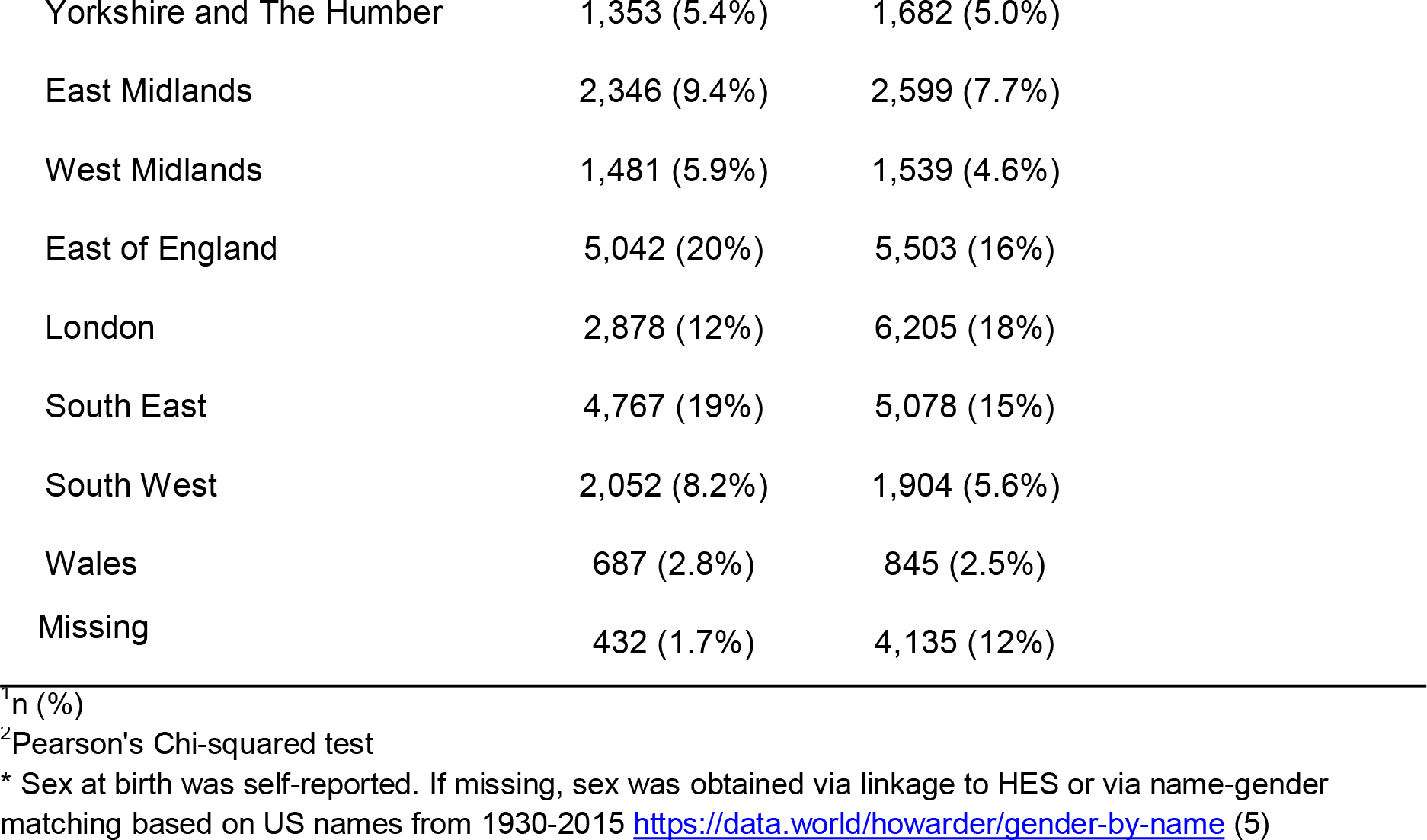
Cohort characteristics stratified by proportion of possible surveys completed (less than or equal to versus more than 75% of surveys completed)

| Characteristic | >=75% of surveys completed<br>N = 24,915<sup>1</sup> | <75% of surveys completed<br>N = 33,713<sup>1</sup> | p-value<sup>2</sup> |
| --- | --- | --- | --- |
| **Age group** |  |  | <0.001 |
| 0-15 | 1,512 (6.1%) | 5,859 (17%) |  |
| 16-24 | 828 (3.3%) | 2,671 (7.9%) |  |
| 25-44 | 2,357 (9.5%) | 9,368 (28%) |  |
| 45-64 | 9,201 (37%) | 10,456 (31%) |  |
| 65+ | 11,017 (44%) | 5,359 (16%) |  |
| **Sex\*** |  |  | <0.001 |
| Male | 11,095 (45%) | 15,179 (45%) |  |
| Female | 13,775 (55%) | 17,758 (53%) |  |
| Missing/Prefer not to say | 45 (0.2%) | 776 (2.3%) |  |
| **Ethnicity** |  |  | <0.001 |
| White British | 21,735 (87%) | 18,746 (56%) |  |
| White Irish | 330 (1.3%) | 341 (1.0%) |  |
| White Other | 1,098 (4.4%) | 1,718 (5.1%) |  |
| Mixed | 312 (1.3%) | 686 (2.0%) |  |
| South Asian | 490 (2.0%) | 2,238 (6.6%) |  |
| Other Asian | 155 (0.6%) | 272 (0.8%) |  |
| Black | 116 (0.5%) | 377 (1.1%) |  |
| Other Ethnicity | 84 (0.3%) | 204 (0.6%) |  |
| Prefer not to say | 53 (0.2%) | 139 (0.4%) |  |
| Missing | 542 (2.2%) | 8,992 (27%) |  |
| **Region** |  |  | <0.001 |
| North East | 1,217 (4.9%) | 1,311 (3.9%) |  |
| North West | 2,660 (11%) | 2,912 (8.6%) |  |

|  |  |  |
| --- | --- | --- |
| Yorkshire and The Humber | 1,353 (5.4%) | 1,682 (5.0%) |
| East Midlands | 2,346 (9.4%) | 2,599 (7.7%) |
| West Midlands | 1,481 (5.9%) | 1,539 (4.6%) |
| East of England | 5,042 (20%) | 5,503 (16%) |
| London | 2,878 (12%) | 6,205 (18%) |
| South East | 4,767 (19%) | 5,078 (15%) |
| South West | 2,052 (8.2%) | 1,904 (5.6%) |
| Wales | 687 (2.8%) | 845 (2.5%) |
| Missing | 432 (1.7%) | 4,135 (12%) |

<sup>1</sup>n (%)
<sup>2</sup>Pearson's Chi-squared test
\* Sex at birth was self-reported. If missing, sex was obtained via linkage to HES or via name-gender matching based on US names from 1930-2015 https://data.world/howarder/gender-by-name (5)

**Figure 1:**
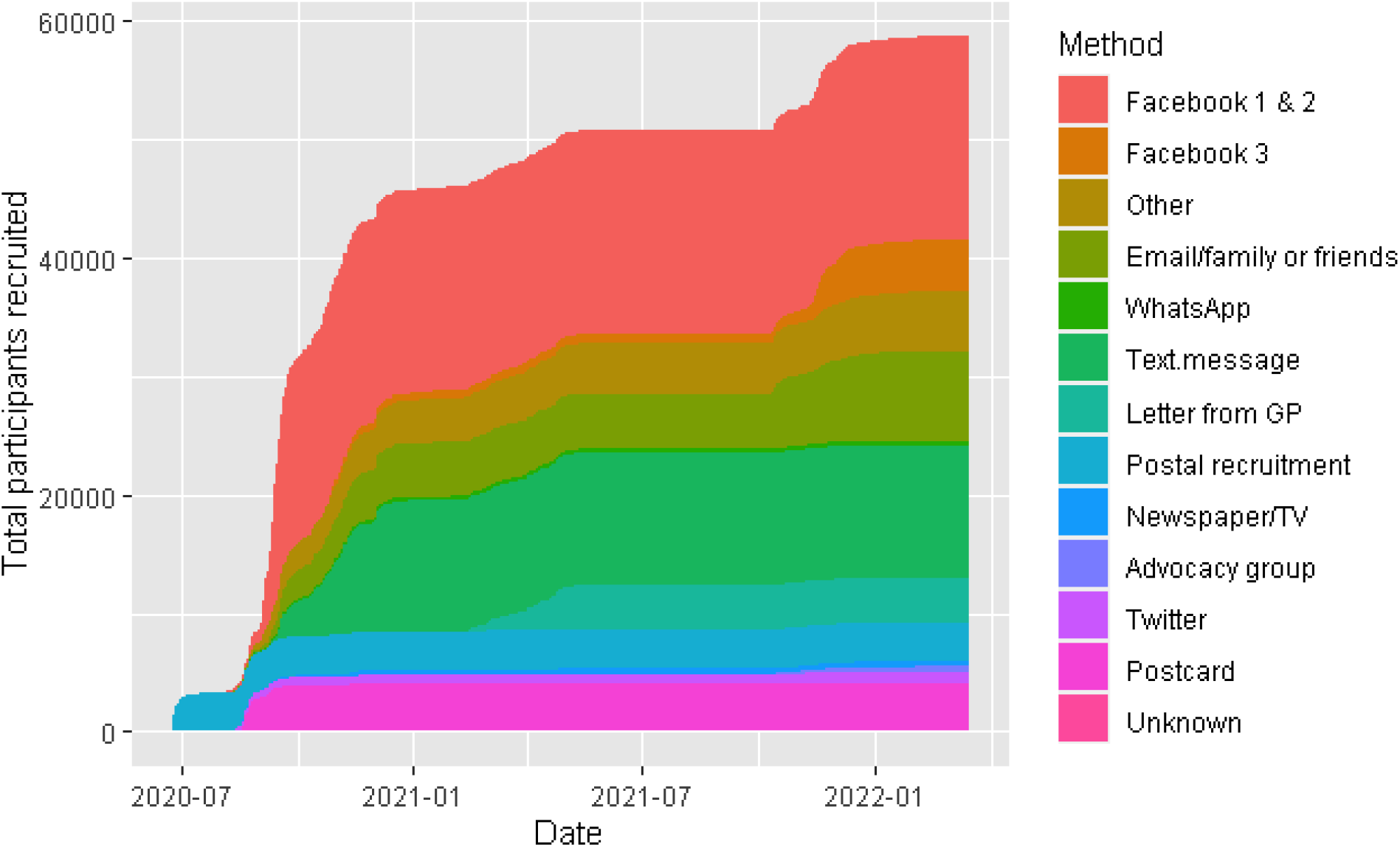
Cumulative participant recruitment by method

**Figure 2:**
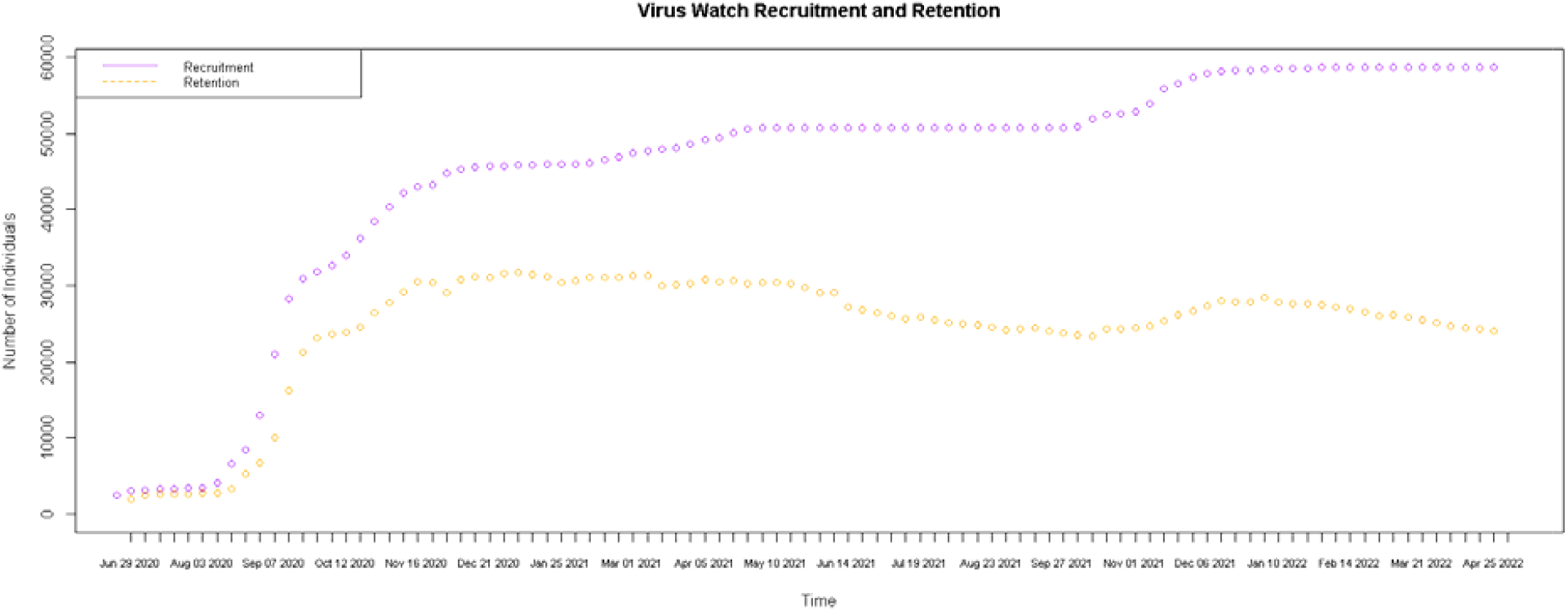
Count of survey completions by week since the start of Virus Watch recruitment (June 2020-May 2022) showing recruitment (total number of participants who completed at least one survey) and retention (total number of participants who completed the latest survey for a given week).

Missing demographics from the baseline survey were requested via a short one-off survey of 5312 study households (n=14,166 participants) in February 2021. This provided 857 (16%) household addresses, the sex of 3,985 (28%) people, and the ethnicity of 3,819 (27%) people.

For records with missing sex data after linkage to NHS Digital data, we assigned gender using the probability of given names being male or female based on US names from 1930-2015 (https://data.world/howarder/gender-by-name) (5). Sex was reported by 49,255 participants, and following gender matching by name, a further 8,552 classifications of sex were inferred. The accuracy of this technique was tested on the 49,255 complete records and found to be 99.82%. Notably, this method fails to account for the minority of individuals who are intersex/other non-binary gender identities and should be interpreted with this in mind.

Data linkage to NIMS for immunisation records up to 23rd December 2021 yielded an additional 11,221 records for Dose 1 of COVID-19 vaccine that were not self-reported, 12,593 for Dose 2, 14,009 for Dose 3, 41 for dose 4 and 12 for Dose 5. We anticipate future data linkage to increase the number of matched missing records for doses 4 & 5. Linkage to the ONS mortality dataset held by NHS Digital, identified 153 participants that had died (up to November 2021) compared to 59 reported deaths by household or family members.

Study participant data were linked to the national ‘Pillar 2 ‘ COVID-19 community testing programme by NHS Digital, which provided an additional 291,067 test results (negative and positive results). We asked participants to self-report only positive test results and the first subsequent negative result via the weekly surveys; consequently only one result was recorded per week per participant. This likely explains the difference between the high number of tests recorded via Pillar 2 compared to our study. Linking to the Second Generation Surveillance System (SGSS) which contains results of testing performed in hospital patients and health and care workers and is held by NHS Digital, provided 5,345 additional test results.

## What has been measured?

Participants completed detailed study questionnaires online via REDCap database tools hosted on the secure UCL Data Safe Haven (6). Demographic, clinical, and socioeconomic data were collected at baseline for each household member, as well as any previous COVID-19 illness. Symptoms, activities, COVID-19 test results and vaccinations were reported in weekly illness surveys and monthly surveys asked questions on a broad range of topics including behaviours, mental health and disability tailored to the pandemic phase. **Table 3** summarises the survey data collected from Virus Watch participants, additional data acquired from linkage via NHS Digital, biological data collected from participants in the laboratory and vaccine efficacy subcohorts and geolocation data from an optional movement tracker app sub-study.

**Table 3.**
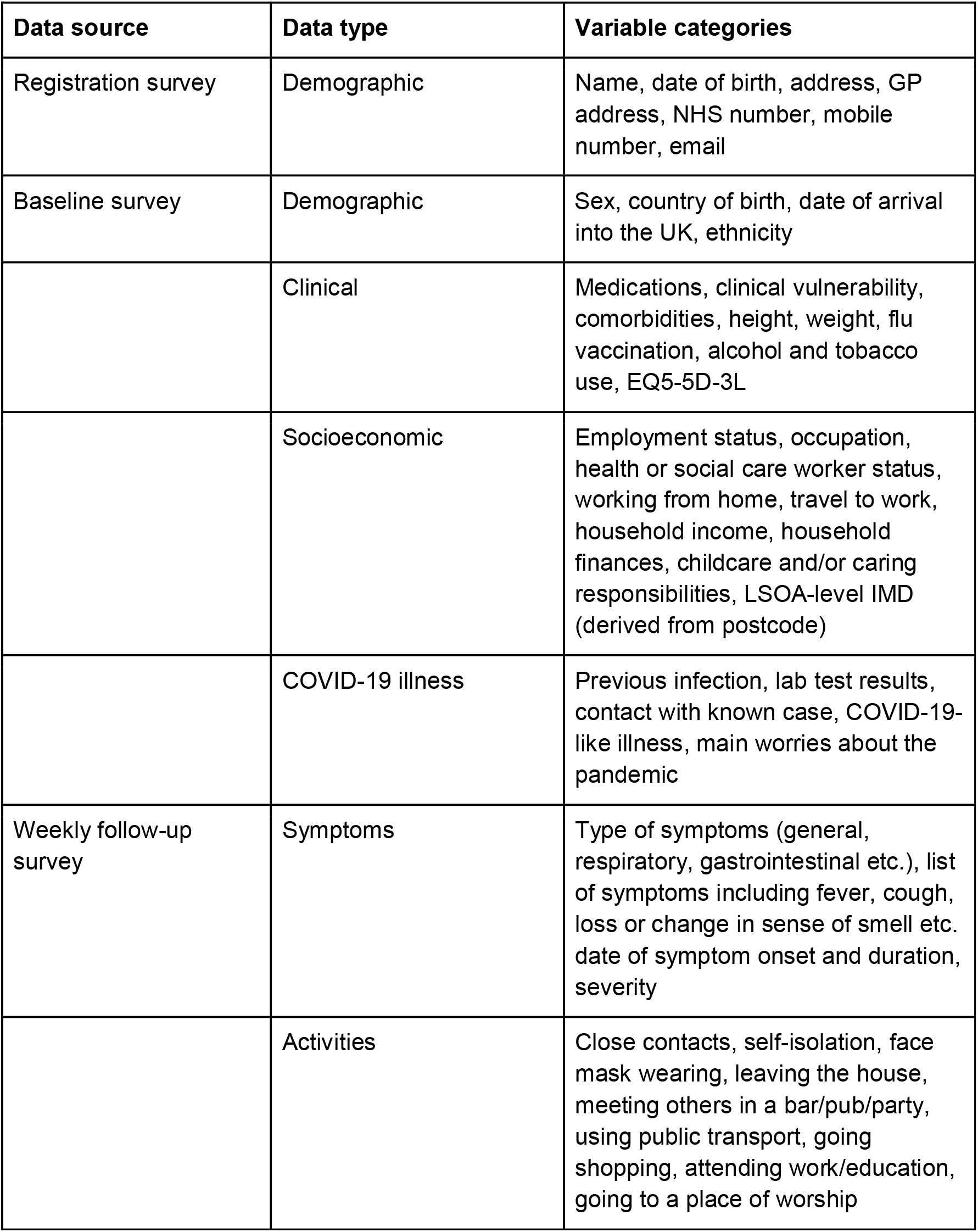
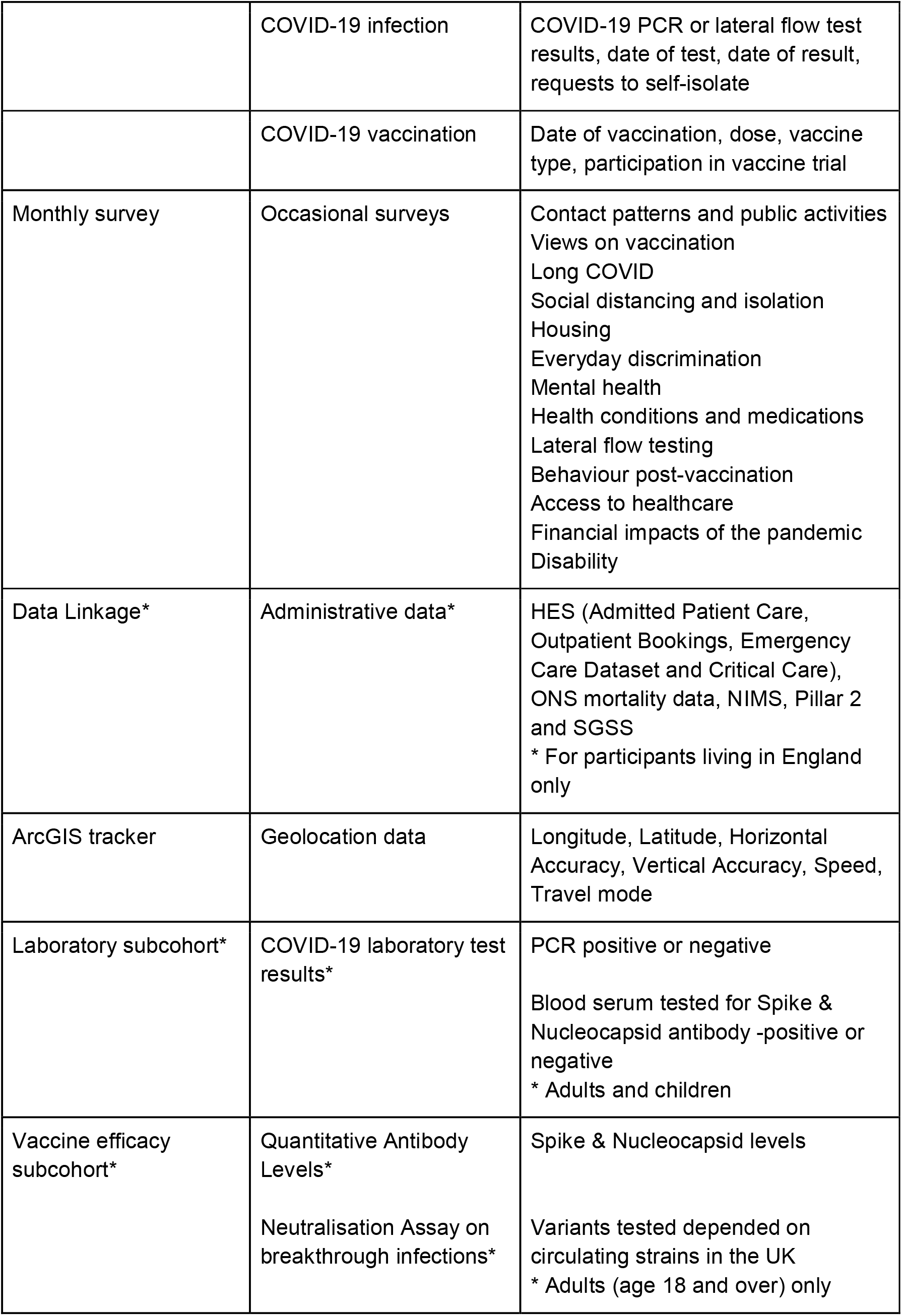
Summary of data items collected from Virus Watch participants, including data source, type of data and top-level categories of variables included.

| Data source | Data type | Variable categories |
| --- | --- | --- |
| Registration survey | Demographic | Name, date of birth, address, GP address, NHS number, mobile number, email |
| Baseline survey | Demographic | Sex, country of birth, date of arrival into the UK, ethnicity |
|  | Clinical | Medications, clinical vulnerability, comorbidities, height, weight, flu vaccination, alcohol and tobacco use, EQ5-5D-3L |
|  | Socioeconomic | Employment status, occupation, health or social care worker status, working from home, travel to work, household income, household finances, childcare and/or caring responsibilities, LSOA-level IMD (derived from postcode) |
|  | COVID-19 illness | Previous infection, lab test results, contact with known case, COVID-19-like illness, main worries about the pandemic |
| Weekly follow-up survey | Symptoms | Type of symptoms (general, respiratory, gastrointestinal etc.), list of symptoms including fever, cough, loss or change in sense of smell etc. date of symptom onset and duration, severity |
|  | Activities | Close contacts, self-isolation, face mask wearing, leaving the house, meeting others in a bar/pub/party, using public transport, going shopping, attending work/education, going to a place of worship |
|  | COVID-19 infection | COVID-19 PCR or lateral flow test results, date of test, date of result, requests to self-isolate |
|  | COVID-19 vaccination | Date of vaccination, dose, vaccine type, participation in vaccine trial |
| Monthly survey | Occasional surveys | Contact patterns and public activities<br>Views on vaccination<br>Long COVID<br>Social distancing and isolation<br>Housing<br>Everyday discrimination<br>Mental health<br>Health conditions and medications<br>Lateral flow testing<br>Behaviour post-vaccination<br>Access to healthcare<br>Financial impacts of the pandemic<br>Disability |
| Data Linkage* | Administrative data* | HES (Admitted Patient Care, Outpatient Bookings, Emergency Care Dataset and Critical Care), ONS mortality data, NIMS, Pillar 2 and SGSS<br>* For participants living in England only |
| ArcGIS tracker | Geolocation data | Longitude, Latitude, Horizontal Accuracy, Vertical Accuracy, Speed, Travel mode |
| Laboratory subcohort* | COVID-19 laboratory test results* | PCR positive or negative<br>Blood serum tested for Spike & Nucleocapsid antibody -positive or negative<br>* Adults and children |
| Vaccine efficacy subcohort* | Quantitative Antibody Levels*<br>Neutralisation Assay on breakthrough infections* | Spike & Nucleocapsid levels<br>Variants tested depended on circulating strains in the UK<br>* Adults (age 18 and over) only |

As of 7th December 2022, 1,619,300 weekly surveys had been submitted with 21,299,348 person days of follow up. 351,751 individual responses to monthly surveys were received, 190,993,995 GPS coordinates. From laboratory substudies we collected 10,974 full serological samples, 107,708 finger prick samples and of these, 4,972 live virus neutralisation activity of capillary microsamples were tested.

## What have we found?

Virus Watch aimed to provide evidence on the transmission and impact of COVID-19 and aimed to estimate key epidemiological measures including: the incidence of PCR-confirmed COVID-19; incidence of hospitalisation among PCR-confirmed COVID-19 cases; incidence of respiratory infection symptoms, including COVID-19 disease case definitions; and secondary household attack rates. Other outcomes of interest included investigating the effectiveness and impact of control measures including testing, isolation, social distancing and vaccine effectiveness against asymptomatic and symptomatic infections.

Early in the pandemic, we took an active decision to avoid duplication of effort in reporting the incidence of infection and hospitalisation once the ONS COVID-19 Infection Survey and the UK Health Security Agency dashboard data were established and chose to focus on investigating: the symptoms of COVID-19 and COVID-19-like illness; risk factors for, and behaviours associated with infections and vaccination; and immunity against COVID-19. We have published a summary of key findings on the study website https://ucl-virus-watch.net/?page_id=1323 and a full list of publications and pre-prints is available at: https://ucl-virus-watch.net/?page_id=1388.

More deprived communities have been disproportionately impacted by the health, social and economic effects of the COVID-19 pandemic. Greater day-to-day exposure to people outside of their household and/or support bubble (e.g. lesser ability to work from home, greater dependence on public transport, etc) may be driving higher infections, hospitalisations and deaths in deprived areas. To explore this we used participant reported data on daily activities during 3 weekly periods in late November 2020, late December 2020 and mid-February 2021 (7). During the final week of November and the December holiday period (23-27 Dec 2020), participants living in more deprived areas were more likely to: leave their house to go to work or school; use public transport; share a car with a non-household member; visit an essential shop; and have close contact with a non-household/support bubble member than participants living in the least deprived areas. Participants living in more deprived areas were not more likely to undertake social and entertainment activities or visit non-essential shops and services. Our findings suggest that differences in essential daily activities are likely to be contributing to higher infection rates in more deprived regions. These differences are likely to reflect circumstances that constrain individual choice, e.g. car ownership, ability to work from home and disposable income. There was no observed difference in activities that are more likely to reflect individual decision making, such as attending non-essential shops or social and entertainment activities.

Work investigating anti-spike SARS-CoV-2 antibody levels among Virus Watch participants who had received COVID-19 vaccines provided key insights into antibody waning and protection against breakthrough infections (8–11). In one analysis following the vaccine rollout in the UK, we measured antibody levels in almost 9000 study participants who had received two doses of ChAdOx-S1 or BNT162b2 vaccine at 3 weeks after the second dose and 20 weeks after the second dose (9). Antibody levels dropped at the same rate for both vaccines, but peak levels were much higher following the BNT162b2 vaccine. We found those with lower antibody levels were at increased risk of infection. We also showed that peak anti-S levels are higher post-booster than post-second dose, but that levels are projected to be similar after six months for BNT162b2 recipients. While peak antibody levels post-second dose were substantially lower for ChAdOx-S1 than BNT162b2 recipients, no differences in post-booster antibody levels by primary course type were observed (**Figure S1**). The magnitude and trajectory of post-booster anti-S response was similar across age groups and by clinical vulnerability status. Higher peak anti-S levels post-booster may partially explain the increased effectiveness of booster vaccination compared to two-dose vaccination against symptomatic infection with the Omicron variant.

## What are the main strengths and weaknesses?

Following up whole households longitudinally, rather than being limited to individuals is a unique strength of Virus Watch. The cohort has data on all ages including children and adolescents and is broadly representative of the UK population on geographic spread and deprivation (**Table 1**), with some exceptions. A particular focus was placed on engaging participants from minority ethnic groups with recruitment methods and research question prioritisation being guided by our study advisory group composed of community leaders, policy experts and charity representatives. Study surveys and data collection methodologies were developed based upon 6 years of experience running large national surveillance cohorts of pandemic and seasonal influenza (12)and we used validated questionnaires wherever feasible (e.g. PHQ-9 and GAD-7 to assess depression and anxiety within the cohort).

The study dataset has been linked to national Pillar 2 testing (PCR & lateral flow testing data through national Test Trace and Isolate Programme), the national vaccine register as well as hospitalisations and deaths. We have collected detailed demographic information, clinical co-morbidity data and have stored serum from participants in the laboratory subcohort at 2 time points, as well as longitudinal serum micro-samples from the vaccine evaluation subcohort. Virus Watch is one of the few longitudinal studies to present quantitative spike and nucleocapsid antibody test data among adults and qualitative (positive/negative) serology among children, together with detailed vaccination information and clinical comorbidities The cohort has several important limitations. Households self-selected into the study after receiving an invitation via multiple routes, biassing the sample towards participants with an interest in COVID-19 and health research. Households with more than six members were not eligible for the study due to the limitations of the REDCap survey infrastructure, and people living in institutional settings such as care homes, university halls of residence and boarding schools were not eligible to participate, limiting the generalisability of findings for these groups. Households also needed to have either a mobile telephone, tablet or computer with an internet connection, a valid email address and at least one household member who could read and respond in English to complete regular surveys.

Several demographic groups are underrepresented in the study. The cohort is older (mean age = 48 years), with a greater proportion of people in the 45–64-year age group when compared to the general population. Some ethnic groups are also under-represented, notably the Black and Other Asian groups (**Table 1**). Our ability to disaggregate data into more granular categories of ethnicity is limited due to the small number of people in these groups enrolled in the study. Retention of participants has decreased significantly over the 2 years the study has been running. As restrictions have been lifted and interest in the pandemic has waned among the general public, around half of the participants enrolled have stopped regularly completing study surveys. Participants who have disengaged are more likely to be younger, from an ethnic minority background and living in London, limiting statistical power and likely biasing analyses using more recent study data.

## Can I get hold of the data? Where can I find out more?

Given the sensitive content in our dataset (information on health, income and household characteristics) for this study, we cannot publish data at the individual level, publicly. We are sharing individual record level data (excluding any data or variables originating from linkage via NHS Digital) on the ONS SRS (DOI: 10.57906/s5f5-nq13). The data are available under restricted access and can be obtained by submitting a request directly to the SRS. We regularly share results and updates on the study via a “Findings so far” section on our website - https://ucl-virus-watch.net/

## Data Availability

https://ons.metadata.works/browser/dataset?id=89201

## Ethics approval

This study has been approved by the Hampstead NHS Health Research Authority Ethics Committee. Ethics approval number – 20/HRA/2320.

This study uses NHS HES (Admitted Patient Care, Outpatient Bookings, Emergency Care Dataset and Critical Care), ONS mortality, Vaccination (NIMS), and COVID-19 testing data (Pillar 2 and SGSS) that were provided within the terms of a data-sharing agreement (DARS-NIC-372269-N8D7Z-V1.6) to the researchers by the Health and Social Care Information Centre (NHS Digital). The data do not belong to the authors and may not be shared by the authors, except in aggregate form for publication. Data can be obtained by submitting a data request through the NHS Digital Data Access Request Service.

## Author contributions

### Contribution Authors

Conceptualisation and study design AH, EF, JK, PH, EN, IC, VL, RAM, TC, AMJ, SM, JG, RG, AR, RWA

Project administration AH, EF, JK, VN, SB, TB, AA, PH, LW, WLEF, CG, PP, MSh, AMDN, EN, MSp, RWA

Data management VN, WLEF, SB, AMDN, CG, TB, RWA

Writing–original draft preparation JK, TB

Writing–review and editing All authors

## List of acronyms

DHSC: Department of Health and Social Care
GP: General Practitioner
HES: Hospital Episode Statistics
IMD: Index of Multiple Deprivation
LCRN: Local Clinical Research Network
MRC: Medical Research Council
NHS: National Health Service
NIHR: National Institute for Health and Care Research
NIMS: National Immunisation Management Service
ONS: Office for National Statistics
PCR: Polymerase Chain Reaction
SGSS: Second Generation Surveillance System
SRS: Secure Research Service
UKRI: UK Research and Innovation

