## Supplementary Materials for "Cohort profile: Virus Watch: Understanding community incidence, symptom profiles, and transmission of COVID-19 in relation to population movement and behaviour"

**Table S1: Recruitment rates by method of recruitment into Virus Watch**

| **Recruitment Method** | **Number Recruited** | **Denominator** | **Recruitment rate** |
| --- | --- | --- | --- |
| Facebook campaign 1&2 (30/07/2020 - 26/10/2021) | 17,156 (29.4%) | 633,844 | 2.71% |
| Facebook campaign 3 (27/10/2021 onwards) | 4338 (7.4%) | 284,729 | 1.52% |
| SMS message from GP | 11,180 (19%) | 799,312 | 1.40% |
| Email / friend family | 7561 (12.9%) | 18,255 | 41.46% |
| Other | 4,986 (8.5%) |  |  |
| Postcard/Flyer drop | 3,913 (6.7%) | 200,000 | 1.96% |
| GP letter targeting minority ethnic population | 3,785 (6.5%) | 91,310 | 4.15% |
| Postal recruitment | 3,289 (5.6%) | 49,120 | 6.70% |
| Twitter | 872 (1.5%) |  |  |
| Clinical patient group | 510 (0.9%) |  |  |
| Newspaper/TV | 423 (0.7%) |  |  |
| Whatsapp | 391 (0.7%) |  |  |
| Unknown | 225 (0.4%) |  |  |
| TOTAL | 58,628 |  |  |

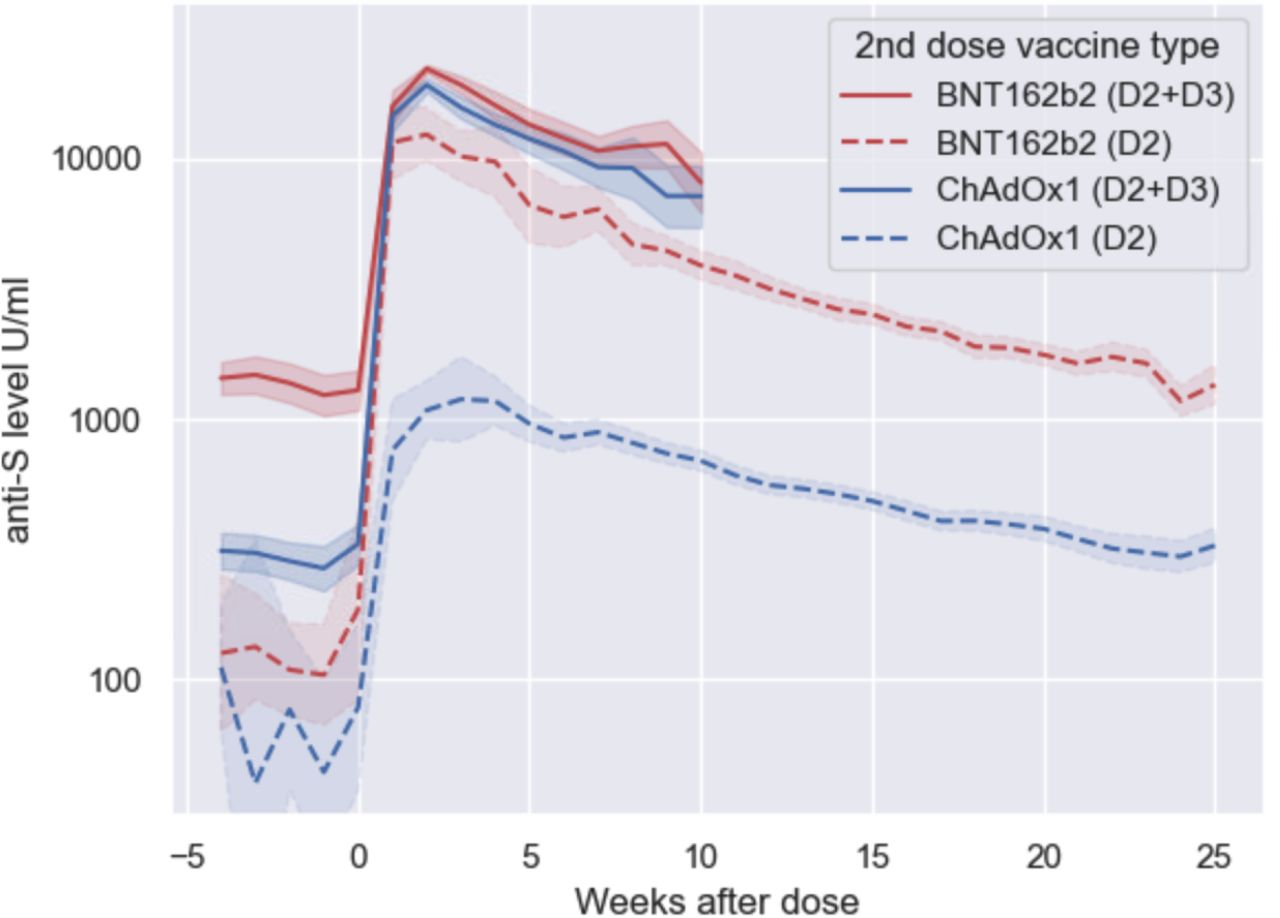

**Figure S1:** Anti-S levels (U/mL) over time since BNT162b2 booster dose (D2+D3) and second vaccine dose (D2) amongst individuals without evidence of prior infection by primary course type.
